# Approaches to defining health facility catchment areas in sub-Saharan Africa

**DOI:** 10.1101/2022.08.18.22278927

**Authors:** Peter M Macharia, Julius N Odhiambo, Eda Mumo, Alex Maina, Emanuele Giorgi, Emelda A Okiro

## Abstract

The geographical area around a health facility characterizing the population that utilizes some or all of its services – a health facility catchment area (HFCA)-, forms the fundamental basis of estimating reliable population denominator for disease mapping and routine healthcare planning. Consequently, the approaches used to delineate the catchment area have a direct impact on the health of a population. To date, there is no systematic literature review documenting different approaches that have been used to define HFCAs while elucidating the implications on derived population denominators. To fill this gap, we systematically reviewed literature and documented approaches that have been used to define HFCA in sub–Saharan Africa (SSA). Simple to complex approaches have been used to define catchment areas in SSA with varying degrees of complexity and limitations in the last four decades. These approaches are mainly driven by lack of geocoded data on the residential address of care seekers and their care-seeking behaviour. To generate closer-to-reality HFCA, for robust disease mapping and healthcare planning, additional data and innovative approaches balancing between model complexity and routine programmatic use are required.

## Introduction

A health facility catchment area (HFCA), also known as a sphere of influence, tributary area, service area or demand field, represents a geographical area around a health facility describing the majority population that uses its services [1,2]. A HFCA is needed to define the catchment population (denominator), which is essential for disease mapping, optimizing timely routine and emergency access, immunization campaigns and vaccination programmes, distribution and allocation of essential health commodities, and planning the location of a new health facility [3–9]. Therefore, knowledge of HFCAs is important for efficient healthcare planning, and resource allocation within a population [10].

Defining a representative HFCA is non-trivial [11]. Its definition is substantially dependent on the availability of geo-positioned residential addresses of patients linked to the sought facility and robust data on their health-seeking behaviour. The healthcare-seeking behaviour is influenced by social-economic, cultural, and religious factors, transport systems, weather patterns, and the characteristics of facilities such as size, services offered, stock-outs, and competition from other health facilities [1,2,6,12]. However, in the majority of sub-Saharan Africa (SSA) and other low-resource settings, such data are not readily available due to limited resources in the context of many competing needs [2]. The problem is more pronounced in rural poor settings where formal address systems are almost non-existent [13]. In addition, privacy concerns may limit the use of precise spatial locations for the residential address of the patients [13,14].

As a result, reliable HFCAs have not been adequately defined by the ministries of Health (MoH) [2,12,15] which hampers routine planning and surveillance [12]. Current attempts have involved the use of the most fundamental data (health facility location and a set of simple auxiliary factors) to define HFCA [6] using a range of simple to complex approaches. However, these approaches are conveniently implemented, disregarding the implications of the defined HFCA on the accuracy of the catchment population and consequences for public service planning. To date, there has been no review of approaches that have been used to define HFCA in SSA to harness the best practices and innovations in defining a closer-to-reality HFCA. Here, we review approaches that have been used to define HFCA in SSA while documenting their pros and cons. We conclude by proposing a pragmatic approach based on the best practices of published literature that can be applied in the SSA context.

## Methods

Our review followed updated guidelines from the Preferred Reporting Items for Systematic Reviews and Meta-Analysis (PRISMA) [16].

### Search strategy

To identify eligible papers, a comprehensive search strategy was developed under the guidance of an information and library expert and a group of spatial epidemiologists. First, we developed a search strategy that leveraged the unique search optimization features and indexing of each of the three electronic databases: PubMed, Scopus and CINHAL. The final literature search was conducted in April 2022. The main search terms were HFCA and its synonyms such as hospital catchment, health service area, facility service area combined with defining/creation/modelling/estimation/delineation/planning and the list of SSA countries. Boolean operators and asterisks were used to optimize the search process. We also screened the bibliography of the selected papers for additional papers. We used Mendeley and Rayyan to serve as bibliographic software for managing references in the review.

### Eligibility criteria

The review sought to identify studies that were closely related to the measurement and conceptualization of HFCA in any SSA country. We did not limit the search by year; therefore, all years were included in the review. We excluded reviews, editorials, and conference presentations but included any relevant studies from their bibliography. We screened the identified studies in three stages: (1) screening by title, (2) screening by abstract, and (3) screening by reading the full text. Two authors independently reviewed all abstracts and full-text formats of the studies while a third author was used to resolve discordances. After screening, data were extracted from the remaining studies.

### Data extraction

An online data extraction form was developed to obtain information about HFCA models and other important study characteristics. These characteristics were 1) bibliographic information, 2) facility level 3) health or study outcome, 4) analytical method used to define HFCA, 5) data needed to define HFCA, 6) sensitivity analysis, 7) and modelling gaps and recommendations that were acknowledged by the authors. Extraction discrepancies were resolved by consensus and by an independent arbitrator.

### Data synthesis

Given the large scope of the review and the heterogeneity of the studies reviewed, a meta-analysis was not appropriate. However, a qualitative synthesis was conducted to identify approaches and methodological commonalities across studies and contexts.

## Result

Overall, we retrieved 808 articles which were exported to the Mendeley reference manager. Studies were screened and excluded by title, abstract, and full text. Studies excluded after full-text review did not explicitly define an approach to model HFCA. Ultimately, 83 peer-reviewed articles met the inclusion criteria (Figure 1). The earliest manuscript was published in 1977 while the majority of the studies (84%) were published after 2008 with 2020-21 contributing 30% of all the studies. The studies varied by geographical scale and scope. Four studies (5%) were conducted across multiple countries whereas 79 studies (95%) were conducted within 21 individual SSA countries. Kenya (16%), Uganda (13%) and Zimbabwe (10%) had the highest number of studies while nine countries, each had at most two studies.

**Figure 1:**
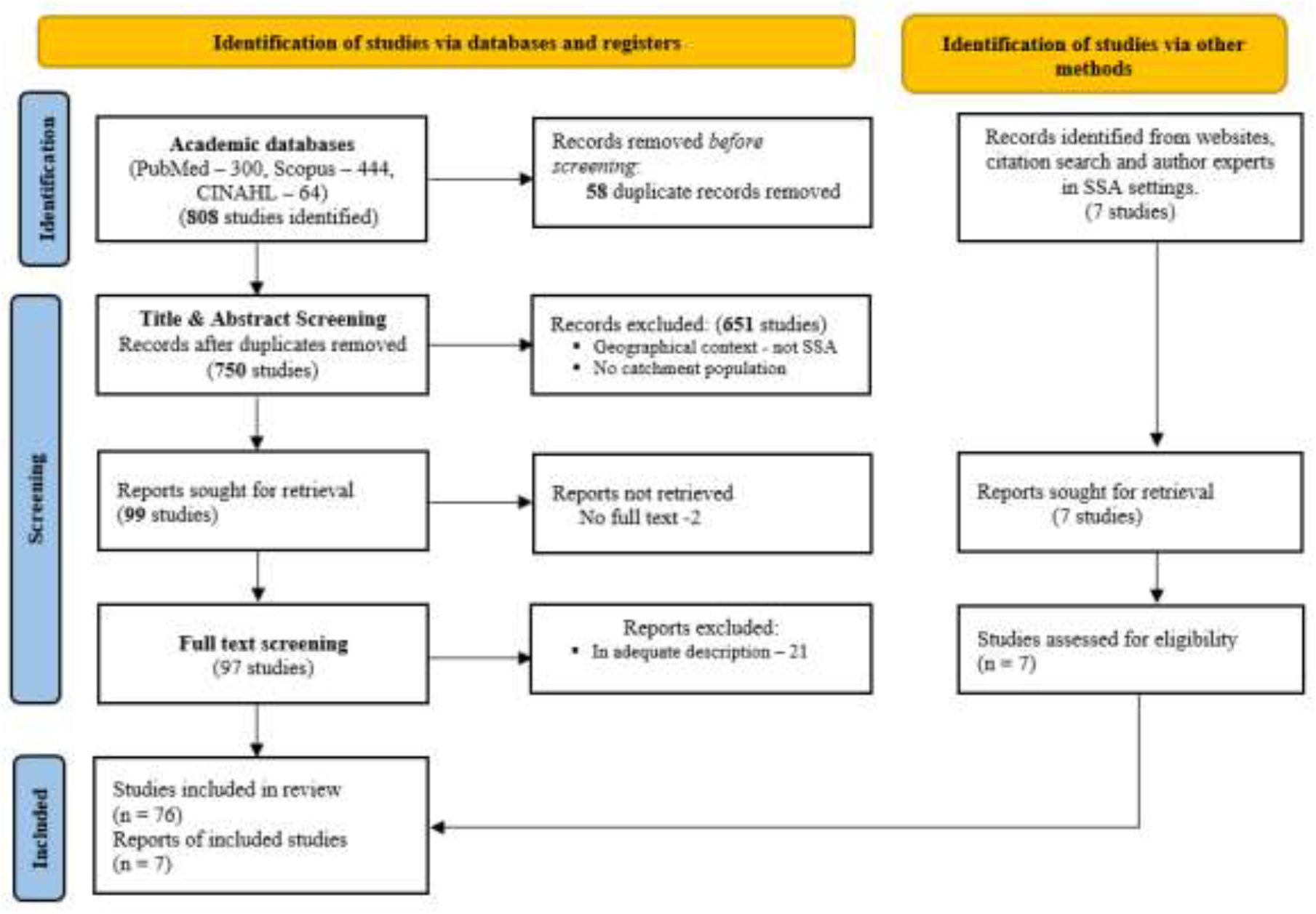
Flowchart for study selection from literature search to data extraction and analyses.

The studies were evenly distributed across the health system hierarchical structure, focusing on either primary (28.9%), secondary (33.7%), or both tiers (37.3%). The main health outcomes across the studies were highly variable. We identified 22 different health outcomes, with malaria (26.5%), HIV (15.7%), healthcare utilization (15.7%), vaccination (6.0%) and maternal and newborn care (6.0%) featuring in 70% of the manuscripts. Table 1 summarises data, threshold used to delineate HFCA, limitations (where indicated) and approaches, that have been used to define HFCA across the last 4 decades in SSA.

**Table 1:**
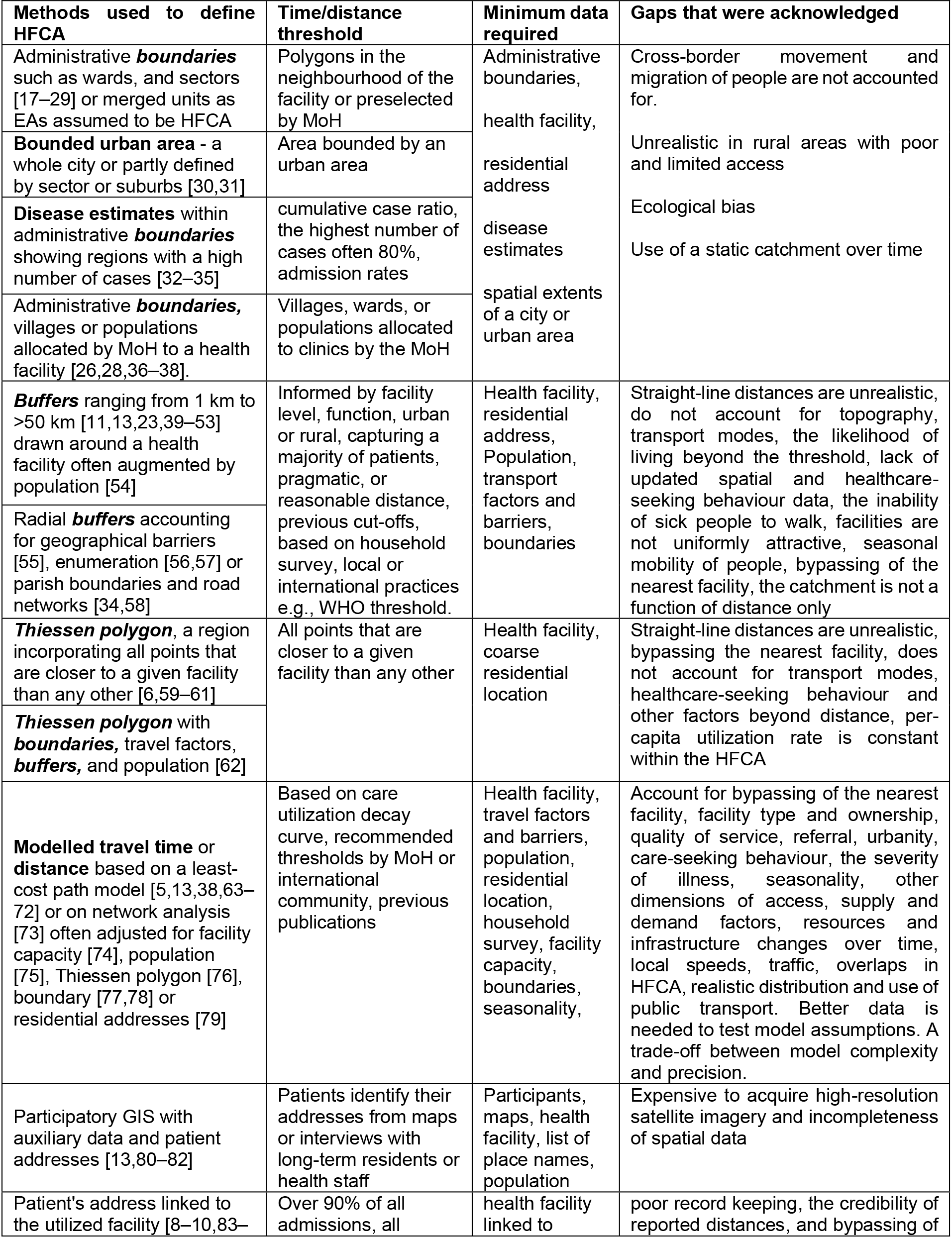
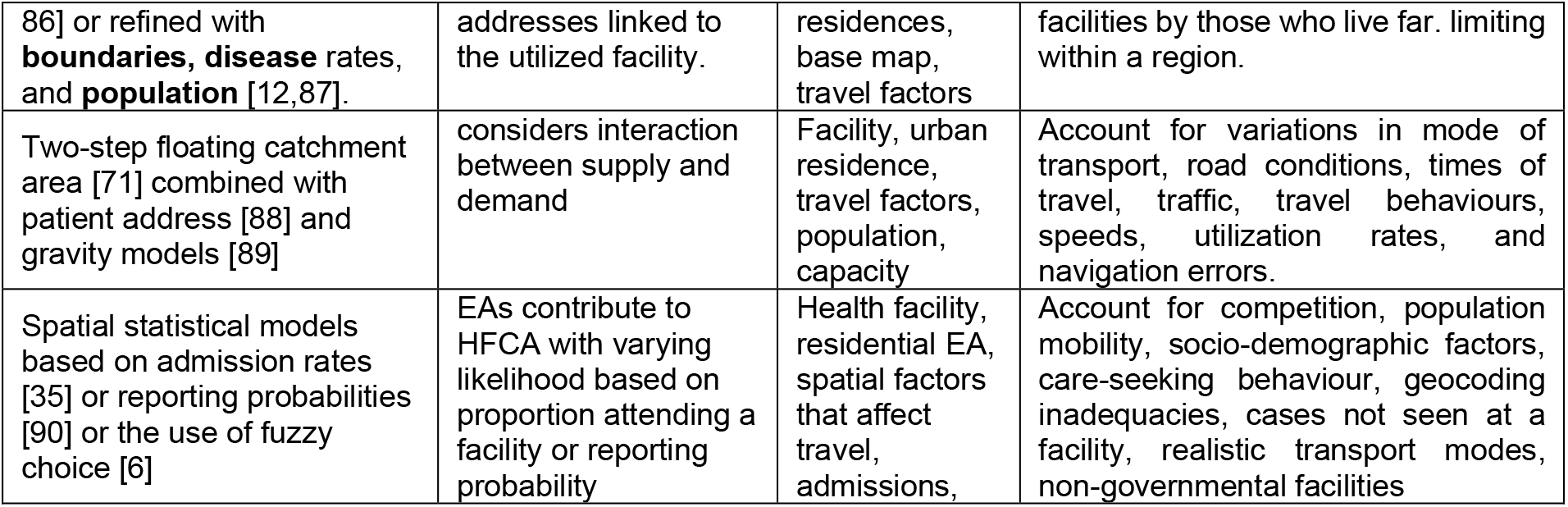
Summary of methods used to generate health facility catchment areas in sub-Saharan Africa including required datasets and limitations of the approaches

The use of subnational administrative boundaries (e.g., wards) to define HFCA was the second most common approach (21 studies). The boundaries of a polygon in which a facility was located and sometimes the neighbouring polygons formed the HFCA. Boundaries were used either independently or in combination with urban areas, disease estimates, population count or allocated by the MoH. The location of the health facility and the administrative boundaries combined with auxiliary datasets were the minimum dataset required. However, this approach ignores cross-border movement, and migration in and out of the catchment over time, particularly in rural areas where alternatives are limited.

Buffers around a health facility defining HFCA require the location of the health facility as the only input. Due to this simplicity, it was the most common approach (24 studies) including those that refined the buffers using population, administrative boundaries, or road networks (Table 1). The buffer size was based on a pragmatic distance that captured most patients or thresholds derived from literature, household surveys, local or international practices, facility level/function, and locality (urban or rural). The use of buffers was criticized because it is based on unrealistic straight-line distances which do not account for topography, transport modes, seasonality, mobility of people, the attractiveness of facilities, the inability of sick people to walk, and documented healthcare-seeking behaviour (such as bypassing the nearest facility). The authors justified the approach given the lack of updated data, especially healthcare-seeking behaviour data.

Closely related to the buffers is the use of Thiessen polygons also known as Voronoi diagrams to define HFCA (4 studies). They define a region incorporating all points that are closer to a given facility than any other facility, have similar data requirements and limitations as the buffers and can be combined with other approaches (Table 1). The need to account for the variable per-capita utilization rate within the HFCA was an additional limitation that was highlighted.

To account for some of the limitations in the use of administrative areas, buffers and Thiessen polygons, in defining HFCA, 20 studies applied a threshold on modelled travel time/distance to define a slightly improved HFCA. Time or distance was modelled through the path of least resistance via network analysis or cost distance surface accounting for transport mode, speeds, travel barriers (game park, reserves, water bodies and forest), travel factors (road network, land cover, topography) and sometimes simplified healthcare seeking behaviour. In some instances, modelled time was adjusted for facility capacity, and population or used in combination with Thiessen polygon, boundary or residential addresses (Table 1). The choice of the threshold was based on previous publications, policy recommendations by MoH or the international community or the use of a utilization decay curve.

Despite accounting for some limitations, drawbacks of modelled travel time/distance to define HFCA exist. Authors recognised the need to better account for care-seeking behaviour (bypassing the nearest facility, severity of illness, other dimensions of access, localised speeds, traffic, weather seasonality, urbanicity, realistic distribution and use of public transport, resources and infrastructure variation over time) supply-side factors (facility type and ownership, quality of services at facilities, referral patterns), and overlapping of two or more HFCA. Further, the authors argued that better data are needed to test model assumptions while balancing the trade-offs between model complexity, precision and routine application.

Albeit minimal, four studies used public participatory Geographical Information System (GIS) approaches. This involved community members and the patients defining HFCA, for example through data collection. Patients would identify their residential addresses from maps presented to them during health facility visits or interviews with long-term residents of an area or health staff to map HFCA. The main requirements were the participants, maps, imagery or a list of place names of the area. The approach was limited given the cost associated with acquiring high-resolution satellite imagery of the area and the incompleteness of existing spatial data.

On the other hand, the use of geocoded patient addresses (9 studies) linked with the health facility provided the most representative catchment area. The patient’s addresses were available at different spatial resolutions and were often refined or combined with boundaries, disease rates, and population. However, poor record keeping, bypassing of facilities, limiting the catchments within a region and the credibility of reported distances were reported as limitations.

Finally, to advance the approaches using modelled travel time, there were six standalone efforts to derive HFCA based on two-step floating catchment area [71,88], gravity models [89], spatial-statistical [35,90] and fuzzy choice models [6]. Mainly, these approaches had residential areas or enumeration areas contributing to HFCA with varying degrees of likelihood based on several factors (Table 1). Despite having some improvements, they did not satisfactorily account for variations in travel (mode of transport and speeds, road conditions, time of travel, traffic conditions, navigation errors), utilization rates and care-seeking behaviour, competition between facilities, population mobility, socio-demographic factors, geocoding inadequacies, cases not seen at a facility and non-governmental facilities.

Across the studies, a range of techniques were implemented as sensitivity analyses for the derived HFCA. These included deriving several HFCA for the same study area while using different; i) methods [23,34,59], ii) assumptions on healthcare-seeking behaviour [38], iii) population thresholds [80], iv) travel speed [71,88], v) radii for the buffer approach [54,73], vi) several teams validating the generated HFCA [81], and vii) using information criterion to select the best statistical model [35]. Finally, AccessMod and ArcMap were the most used software to derive HFCA. Other software included QGIS, Google Earth, GeoDa, Epi Info, R, STATA, FoxPro, and SAS

## Discussion

The review has outlined approaches that have been used to define HFCA in SSA, a largely resource and data-constrained region. These approaches either rely on or are associated with techniques of defining geographical access as summarised in Ouma et al 2020 [91]. Overall, in SSA, there is a scarcity of geocoded data on patients’ residential addresses linked with the facility where care was sought which is the gold standard in defining a HFCA (Table 2). As a result, only six studies utilized such data [8–10,12,83–87], while six other studies either relied on MoH-derived HFCA [26,28,36] or used participatory GIS to collect data needed to delineate spatial extents of HFCAs [13,80–82]. The rest of the approaches used a variety of methods, with varying degrees of representativeness to delineate HFCA

**Table 2:** Choice of method in generating health facility catchment areas in sub-Saharan Africa and low resource settings

| Proposed level | Approach | Notes |
| --- | --- | --- |
| Level 3: Least appropriate | Thiessen polygons, administrative boundaries, and buffers | Oversimplified assumptions which are unrealistic in terms of healthcare-seeking behaviour and health system characteristics. Thus, should be rarely used unless results are aggregated to large subnational units |
| Level 2: Moderately appropriate | Modelled travel time and distance while accounting for key factors and balancing between model complexity and programme use. | Should robustly account for healthcare-seeking behaviour, realistic transport systems, demand, and supply side of a health system. |
| Level 1: Most appropriate | Geocoded residential address of a patient linked to the utilized health facility at a high spatial resolution | High spatial resolution patient residential addresses with their journey experiences and outcome within the health system should be well documented |

Three commonly used approaches; administrative boundaries, buffers, and Thiessen polygons are limited because they oversimplify socio-demographic, epidemiological and health-seeking characteristics of communities when deriving HFCA (Table 1). These inadequate approaches will thus result in a non-representative catchment population and therefore, their use should be discouraged (Table 2). However, these approaches might be useful for applications that aggregate results to large subnational units. For example, a catchment derived using Thiessen polygons, but results presented at a district level. On the other hand, approaches based on travel time, gravity, and spatial statistical models while useful, also still require novel extensions to deal with their shortcomings (Table 1) to push the frontier to the next level. The advances should be made widely accessible at the programmatic level for routine use.

The key aspects that should be considered to open up a new avenue for HFCA definitions are cross-cross-border movement and overlapping catchments, mobility of patients, realistic travel times (that account for weather seasonality, transport modes within the public and private sector, localised speeds, road conditions, traffic, time of journey, navigation errors), competition between facilities, health-seeking behaviour (bypassing of the nearest facility, socio-demographic factors, severity of illness, cases not seen at a facility), facility characteristics (type and ownership, quality of service), residence (urban or rural) and referral patterns.

To account for these aspects, better data will be needed. This will also aid in testing model assumptions, and deal with the perennial incompleteness of spatial data, poor record keeping and geocoding inadequacies [92]. With the advancement in data science (such as machine learning) and data collection techniques (such as remote sensing), a range of climatic and environmental data (e.g., land use, rainfall patterns), road networks and traffic patterns can now be easily collected [93–95]. Increasingly available household surveys and routine data will be valuable for tracking utilization rates in the population to derive better thresholds for different health outcomes and contexts. Further, the use of mobile phones has become ubiquitous across the globe and can be harnessed to record geographical location information, especially in SSA to improve HFCA definition [96]. However, privacy and data protection concerns will need to be addressed when utilizing data from mobile phones. This is also a challenge affecting sharing of patients’ addresses and locations of service providers in the routine health information systems in SSA.

The travel time or distance thresholds that patients can travel are critical in the delineation of HFCA irrespective of the complexity of the approach. The threshold varies depending on the local context, health condition, severity of illness, and services offered at a facility. The use of healthcare utilization data for a particular outcome to create a decay curve or medical relevant thresholds is more useful than random and generalized thresholds. It is probably the use of random thresholds, cross-border movements, and simplified approaches (administrative boundaries, Thiessen polygons or buffers) that may have led to health coverage exceeding 100% at the facility level in recent DHIS2 analyses [97].

We, therefore, propose three levels when choosing an approach to delineate HFCA guided by the data availability and study objectives (Table 13.2). *Level 1* is the most appropriate approach where HFCA can be defined unambiguously. It will require patients’ addresses to be geocoded and linked with the service provider where care was sought and where possible to harness recent technologies to collect these data. The second level (*Level 2*) is based on travelled time or distance but requires innovative methods to deal with the outlined key shortcomings. Finally, *level 3*, is the least recommended and its use is discouraged due to unrealistic assumptions.

Further MoH-derived HFCAs should be available across countries in SSA as a fundamental baseline for healthcare planning. However, limited studies referenced the use of MoH-derived HFCA, which may imply the absence of guidelines within MoH on defining robust HFCA. This may be attributed to poor documentation, or that the role of HFCA is under-appreciated. In this line, though at nascent stages, is a promising initiative aiming to create a system that enables MoH and stakeholders to define, create and manage their HFCAs [98].

Much of SSA and other low-resource countries are currently striving to achieve the ambitious targets within the Sustainable Development Goals (SDGs) framework by 2030. The SDG mantra of *leaving no one behind, and reaching those farthest, behind, first* would require estimating the populations in need of essential health services and defining health care coverage gaps at the HFCA level for targeted resource location. This will be essential for universal health coverage (UHC) to ensure that all people have access to the health services they need, when and where they need them, without financial hardship. Therefore, the role of accurate HFCA is timely and cannot be ignored as *a catalyst for health development in SSA*. In addition, the concept of a catchment area extends beyond health facilities and similar cases (limitations and requirements) may be advanced for catchment areas related to schools [4], community health workers, and vaccination posts among other service delivery points [2].

The review should be interpreted while considering several limitations. The literature search was limited to studies published in English. Secondly, given the vast nature of grey literature, some insights on HFCA in SSA might have been missed and our findings can only be applied to SSA countries or similar contexts. Despite these limitations, the review shows that most of the studies derived HFCA using simplified approaches due to a lack of appropriate data. To move the frontier of HFCA to the next level, the majority of the limitations that were acknowledged should be accounted for to derive closer-to-reality HFCA for robust catchment populations (denominator) for healthcare planning.

## Data Availability

All data produced in the present work are contained in the manuscript

## Declarations

### Availability of data and materials

All data relevant to the study are included in the article. The data supporting conclusions made in this review are available in the detailed reference list.

### Ethics approval

This study used secondary data only, all publicly available and can be accessed from reference list

### Patient consent for publication

Not required

## References

1 Iyun F. Hospital service areas in Ibadan city. Soc Sci Med 1983;17:601–16. doi:10.1016/0277-9536(83)90304-0

2 Macharia PM, Ray N, Giorgi E, et al. Defining service catchment areas in low-resource settings. BMJ Global Health. 2021;6. doi:10.1136/bmjgh-2021-006381

3 Sturrock HJ, Cohen JM, Keil P, et al. Fine-scale malaria risk mapping from routine aggregated case data. Malaria Journal 2014;13. doi:10.1186/1475-2875-13-421

4 Macharia PM, Ray N, Gitonga CW, et al. Combining school-catchment area models with geostatistical models for analysing school survey data from low-resource settings: Inferential benefits and limitations. Spatial Statistics 2022;51:100679. doi:10.1016/j.spasta.2022.100679

5 Macharia PM, Odera PA, Snow RW, et al. Spatial models for the rational allocation of routinely distributed bed nets to public health facilities in Western Kenya. Malaria Journal 2017;16. doi:10.1186/s12936-017-2009-3

6 Gething PW, Noor AM, Zurovac D, et al. Empirical modelling of government health service use by children with fevers in Kenya. Acta Tropica 2004;91:227–37. doi:10.1016/j.actatropica.2004.05.002

7 Alegana VA, Okiro EA, Snow RW. Routine data for malaria morbidity estimation in Africa: Challenges and prospects. BMC Medicine 2020;18. doi:10.1186/s12916-020-01593-y

8 Airey T. The impact of road construction on the spatial characteristics of hospital utilization in the Meru district of Kenya. Sm Sci Med 1992;34:1135–46.

9 Criel B, Macq J, Bossyns P, et al. A coverage plan for health centres in Murewa District in Zimbabwe: an example of action research. Tropical Medicine and International Health 1996;1:699–709.

10 Kloos H. Utilization of selected hospitals, health centres and health stations in central, southern and western Ethiopia. Soc Sci Med 1990;31:101–14. doi:10.1016/0277-9536(90)90052-t

11 Okiro EA, Bitira D, Mbabazi G, et al. Increasing malaria hospital admissions in Uganda between 1999 and 2009. BMC Medicine 2011;9. doi:10.1186/1741-7015-9-37

12 Okiring J, Epstein A, Namuganga JF, et al. Relationships between test positivity rate, total laboratory confirmed cases of malaria, and malaria incidence in high burden settings of Uganda: an ecological analysis. Malaria Journal 2021;20. doi:10.1186/s12936-021-03584-7

13 Stresman GH, Stevenson JC, Owaga C, et al. Validation of three geolocation strategies for health-facility attendees for research and public health surveillance in a rural setting in western Kenya. Epidemiology and Infection 2014;142:1978–89. doi:10.1017/S0950268814000946

14 Warren JL, Perez-Heydrich C, Burgert CR, et al. Influence of Demographic and Health Survey Point Displacements on Distance-Based Analyses. Spatial Demography 2016;4. doi:10.1007/s40980-015-0014-0

15 CartONG. The challenge of localizing SDGs: CartONG’s data collaborative experience in DRC. 2019.https://cartong.org/news/challenge-localizing-sdgs-data-collaborative-drc (accessed 3 Aug 2022).

16 Page MJ, McKenzie JE, Bossuyt PM, et al. The PRISMA 2020 statement: An updated guideline for reporting systematic reviews. The BMJ. 2021;372. doi:10.1136/bmj.n71

17 McCoy SI, Fahey C, Buzdugan R, et al. Targeting elimination of mother-to-child HIV transmission efforts using geospatial analysis of mother-to-child HIV transmission in Zimbabwe. AIDS 2016;30:1829–37. doi:10.1097/QAD.0000000000001127

18 Chipwaza B, Sumaye RD. High malaria parasitemia among outpatient febrile children in low endemic area, East-Central Tanzania in 2013. BMC Research Notes 2020;13. doi:10.1186/s13104-020-05092-4

19 Emmanuel OW, Samuel AA, Helen KL. Determinants of childhood vaccination completion at a peri-urban hospital in Kenya, December 2013 - January 2014: a case control study. Pan Afr Med J 2015;20:277. doi:10.11604/pamj.2015.20.277.5664

20 Kigozi SP, Kigozi RN, Sserwanga A, et al. Malaria burden through routine reporting: Relationship between incidence and test positivity rates. American Journal of Tropical Medicine and Hygiene 2019;101:137–47. doi:10.4269/ajtmh.18-0901

21 Krumkamp R, Schwarz NG, Sarpong N, et al. Extrapolating respiratory tract infection incidences to a rural area of Ghana using a probability model for hospital attendance. International Journal of Infectious Diseases 2012;16. doi:10.1016/j.ijid.2012.02.003

22 Mugambe RK, Yakubu H, Wafula ST, et al. Factors associated with health facility deliveries among mothers living in hospital catchment areas in Rukungiri and Kanungu districts, Uganda. BMC Pregnancy and Childbirth 2021;21. doi:10.1186/s12884-021-03789-3

23 Peters MA, Mohan D, Naphini P, et al. Linking household surveys and facility assessments: a comparison of geospatial methods using nationally representative data from Malawi. Population Health Metrics 2020;18. doi:10.1186/s12963-020-00242-z

24 Poletti P, Parlamento S, Fayyisaa T, et al. The hidden burden of measles in Ethiopia: How distance to hospital shapes the disease mortality rate. BMC Medicine 2018;16. doi:10.1186/s12916-018-1171-y

25 Sudhof L, Amoroso C, Barebwanuwe P, et al. Local use of geographic information systems to improve data utilisation and health services: Mapping caesarean section coverage in rural Rwanda. Tropical Medicine and International Health 2013;18:18–26. doi:10.1111/tmi.12016

26 Buzdugan R, McCoy SI, Watadzaushe C, et al. Evaluating the impact of Zimbabwe’s prevention of mother-to-child HIV transmission program: Population-level estimates of HIV-free infant survival pre-option A. PLoS ONE 2015;10. doi:10.1371/journal.pone.0134571

27 Koyuncu A, Dufour MSK, McCoy SI, et al. Protocol for the evaluation of the population-level impact of Zimbabwe’s prevention of mother-to-child HIV transmission program option B+: A community based serial cross-sectional study 11 Medical and Health Sciences 1117 Public Health and Health Services. BMC Pregnancy and Childbirth 2019;19. doi:10.1186/s12884-018-2146-x

28 Buzdugan R, Dufour MSK, McCoy SI, et al. Option A improved HIV-free infant survival and mother to child HIV transmission at 9-18 months in Zimbabwe. AIDS 2016;30:1655–62. doi:10.1097/QAD.0000000000001111

29 Marks F, von Kalckreuth V, Aaby P, et al. Incidence of invasive salmonella disease in sub-Saharan Africa: a multicentre population-based surveillance study. The Lancet Global Health 2017;5:e310–23. doi:10.1016/S2214-109X(17)30022-0

30 Bertolote JM, Fleischmann A, de Leo D, et al. Suicide attempts, plans, and ideation in culturally diverse sites: The WHO SUPRE-MISS community survey. Psychological Medicine 2005;35:1457–65. doi:10.1017/S0033291705005404

31 Mboup A, ehanzin LB, Gu edou FA, et al. Early antiretroviral therapy and daily pre-exposure prophylaxis for HIV prevention among female sex workers in Cotonou, Benin: a prospective observational demonstration study. Published Online First: 2018. doi:10.1002/jia2.25208/full

32 Beyene AD, Kebede F, Mammo BM, et al. The implementation and impact of a pilot hydrocele surgery camp for LF-endemic communities in Ethiopia. PLoS Neglected Tropical Diseases 2021;15. doi:10.1371/journal.pntd.0009403

33 Centers for Disease Control and Prevention (CDC). Estimating Meningitis Hospitalization Rates for Sentinel Hospitals Conducting Invasive Bacterial Vaccine-Preventable Diseases Surveillance. Morbidity and Mortality Weekly Report 2013;62:810–2.

34 Zinszer K, Charland K, Kigozi R, et al. Determining health-care facility catchment areas in Uganda using data on malaria-related visits. Bull World Health Organ 2014;92:178–86. doi:10.2471/BLT.13.125260

35 Alegana VA, Khazenzi C, Akech SO, et al. Estimating hospital catchments from in-patient admission records: a spatial statistical approach applied to malaria. Scientific Reports 2020;10. doi:10.1038/s41598-020-58284-0

36 Kruk ME, Hermosilla S, Larson E, et al. Bypassing primary care clinics for childbirth: a crosssectional study in the Pwani region, United Republic of Tanzania. Bull World Health Organ 2014;92:246–53. doi:10.2471/BLT.13.126417

37 Paireau J, Maïnassara HB, Jusot J-F, et al. Spatio-Temporal Factors Associated with Meningococcal Meningitis Annual Incidence at the Health Centre Level in Niger, 2004-2010. PLoS Neglected Tropical Diseases 2014;8. doi:10.1371/journal.pntd.0002899

38 Arambepola R, Keddie SH, Collins EL, et al. Spatiotemporal mapping of malaria prevalence in Madagascar using routine surveillance and health survey data. Scientific Reports 2020;10. doi:10.1038/s41598-020-75189-0

39 Atela M, Bakibinga P, Ettarh R, et al. Strengthening health system governance using health facility service charters: A mixed methods assessment of community experiences and perceptions in a district in Kenya. BMC Health Services Research 2015;15. doi:10.1186/s12913-015-1204-6

40 Gonese E, Dzangare J, Gregson S, et al. Comparison of HIV prevalence estimates for Zimbabwe from antenatal clinic surveillance (2006) and the 2005-06 Zimbabwe demographic and health survey. PLoS ONE 2010;5. doi:10.1371/journal.pone.0013819

41 Hensen B, Lewis JJ, Schaap A, et al. Frequency of HIV-testing and factors associated with multiple lifetime HIV-testing among a rural population of Zambian men. BMC Public Health 2015;15:960. doi:10.1186/s12889-015-2259-3

42 Hensen B, Lewis JJ, Schaap A, et al. Factors associated with HIV-testing and acceptance of an offer of home-based testing by men in rural Zambia. AIDS Behav 2015;19:492–504. doi:10.1007/s10461-014-0866-0

43 Lori JR, Boyd CJ, Munro-Kramer ML, et al. Characteristics of maternity waiting homes and the women who use them: Findings from a baseline cross-sectional household survey among SMGL-supported districts in Zambia. PLoS ONE 2018;13. doi:10.1371/journal.pone.0209815

44 Lori JR, Perosky J, Munro-Kramer ML, et al. Maternity waiting homes as part of a comprehensive approach to maternal and newborn care: A cross-sectional survey. BMC Pregnancy and Childbirth. 2019;19. doi:10.1186/s12884-019-2384-6

45 Njuki R, Obare F, Warren C, et al. Community experiences and perceptions of reproductive health vouchers in Kenya. BMC Public Health 2013;13. doi:10.1186/1471-2458-13-660

46 Ochoa-Moreno I, Bautista-Arredondo S, McCoy SI, et al. Costs and economies of scale in the accelerated program for prevention of mother-to-child transmission of HIV in Zimbabwe. PLoS ONE 2020;15. doi:10.1371/journal.pone.0231527

47 Parker RK, Dawsey SM, Abnet CC, et al. Frequent occurrence of esophageal cancer in young people in western Kenya. Diseases of the Esophagus 2010;23:128–35. doi:10.1111/j.1442-2050.2009.00977.x

48 Walker G, Gish O. Inequality in the distribution and differential utilization of health services: a Botswana case study. J Trop Med Hyg 1977;80:238–43.

49 Kamau A, Mogeni P, Okiro EA, et al. A systematic review of changing malaria disease burden in sub-Saharan Africa since 2000: Comparing model predictions and empirical observations. BMC Medicine 2020;18. doi:10.1186/s12916-020-01559-0

50 Ashiagbor G, Ofori-Asenso R, Forkuo EK, et al. Measures of geographic accessibility to health care in the Ashanti Region of Ghana. Sci Afr 2020;9. doi:10.1016/j.sciaf.2020.e00453

51 Chakraborty NM, Mbondo M, Wanderi J. Evaluating the impact of social franchising on family planning use in Kenya. J Health Popul Nutr 2016;35:19. doi:10.1186/s41043-016-0056-y

52 Montana LS, Mishra V, Hong R. Comparison of HIV prevalence estimates from antenatal care surveillance and population-based surveys in sub-Saharan Africa. Sexually Transmitted Infections 2008;84. doi:10.1136/sti.2008.030106

53 Musinguzi J, Kirungi W, Opio A, et al. Comparison of HIV Prevalence Estimates From Sentinel Surveillance and a National Population-Based Survey in Uganda, 2004-2005. 2009. www.jaids.com

54 Haidari LA, Brown ST, Constenla D, et al. Geospatial planning and the resulting economic impact of human papillomavirus vaccine introduction in Mozambique. Sexually Transmitted Diseases 2017;44:222–6. doi:10.1097/OLQ.0000000000000574

55 Doherty J, Rispel L, Webb N. Developing a plan for primary health care facilities in Soweto, South Africa. Part II: Applying locational criteria. Health Policy & Planning 1996;11:394–405. doi:heapol/11.4.394

56 Kamau A, Mtanje G, Mataza C, et al. Malaria infection, disease and mortality among children and adults on the coast of Kenya. Malaria Journal 2020;19. doi:10.1186/s12936-020-03286-6

57 Kamau A, Mtanje G, Mataza C, et al. The relationship between facility-based malaria test positivity rate and communitybased parasite prevalence. PLoS ONE 2020;15. doi:10.1371/journal.pone.0240058

58 Mpimbaza A, Walemwa R, Kapisi J, et al. The age-specific incidence of hospitalized paediatric malaria in Uganda. BMC Infectious Diseases 2020;20. doi:10.1186/s12879-020-05215-z

59 Farber SH, Vissoci JRN, Tran TM, et al. Geospatial Analysis of Unmet Surgical Need in Uganda: An Analysis of SOSAS Survey Data. World Journal of Surgery 2017;41:353–63. doi:10.1007/s00268-016-3689-5

60 Kundrick A, Huang Z, Carran S, et al. Sub-national variation in measles vaccine coverage and outbreak risk: A case study from a 2010 outbreak in Malawi. BMC Public Health 2018;18. doi:10.1186/s12889-018-5628-x

61 Pattnaik A, Mohan D, Tsui A, et al. The aggregate effect of implementation strength of family planning programs on modern contraceptive use at the health systems level in rural Malawi. PLoS ONE 2021;16. doi:10.1371/journal.pone.0232504

62 Okiro EA, Kazembe LN, Kabaria CW, et al. Childhood Malaria Admission Rates to Four Hospitals in Malawi between 2000 and 2010. PLoS ONE 2013;8. doi:10.1371/journal.pone.0062214

63 Alegana VA, Wright JA, Pentrina U, et al. Spatial modelling of healthcare utilisation for treatment of fever in Namibia. International Journal of Health Geographics 2012;11. doi:10.1186/1476-072X-11-6

64 Bailey PE, Keyes EB, Parker C, et al. Using a GIS to model interventions to strengthen the emergency referral system for maternal and newborn health in Ethiopia. International Journal of Gynecology and Obstetrics 2011;115:300–9. doi:10.1016/j.ijgo.2011.09.004

65 Brunie A, MacCarthy J, Mulligan B, et al. Practical implications of policy guidelines: A gis model of the deployment of community health volunteers in madagascar. Global Health Science and Practice 2020;8:466–77. doi:10.9745/GHSP-D-19-00421

66 el Vilaly MA salam, Jones MA, Stankey MC, et al. Access to paediatric surgery: the geography of inequality in Nigeria. BMJ Global Health 2021;6:e006025. doi:10.1136/bmjgh-2021-006025

67 Epstein A, Namuganga JF, Kamya E v, et al. Estimating malaria incidence from routine health facility-based surveillance data in Uganda. Malaria Journal 2020;19. doi:10.1186/s12936-020-03514-z

68 Palk L, Okano JT, Dullie L, et al. Travel time to health-care facilities, mode of transportation, and HIV elimination in Malawi: a geospatial modelling analysis. The Lancet Global Health 2020;8:e1555–64. doi:10.1016/S2214-109X(20)30351-X

69 Ray N, Ebener S. AccessMod 3.0: Computing geographic coverage and accessibility to health care services using anisotropic movemen of patients. International Journal of Health Geographics 2008;7. doi:10.1186/1476-072X-7-63

70 Sturrock HJ, Cohen JM, Keil P, et al. Fine-scale malaria risk mapping from routine aggregated case data. Malaria Journal 2014;13. doi:10.1186/1475-2875-13-421

71 Tansley G, Stewart B, Zakariah A, et al. Population-level Spatial Access to Prehospital Care by the National Ambulance Service in Ghana. Prehospital Emergency Care 2016;20:768–75. doi:10.3109/10903127.2016.1164775

72 Ouma PO, Agutu NO, Snow RW, et al. Univariate and multivariate spatial models of health facility utilisation for childhood fevers in an area on the coast of Kenya. International Journal of Health Geographics 2017;16:34. doi:10.1186/s12942-017-0107-7

73 Tansley G, Schuurman N, Amram O, et al. Spatial access to emergency services in low- and middle-income countries: A GIS-based analysis. PLoS ONE 2015;10. doi:10.1371/journal.pone.0141113

74 Aoun N, Matsuda H, Sekiyama M. Geographical accessibility to healthcare and malnutrition in Rwanda. Social Science and Medicine 2015;130:135–45. doi:10.1016/j.socscimed.2015.02.004

75 Huerta Munoz U, Källestål C. Geographical accessibility and spatial coverage modeling of the primary health care network in the Western Province of Rwanda. International Journal of Health Geographics 2012;11. doi:10.1186/1476-072X-11-40

76 Kigozi SP, Kigozi RN, Sebuguzi CM, et al. Spatial-temporal patterns of malaria incidence in Uganda using HMIS data from 2015 to 2019. BMC Public Health 2020;20. doi:10.1186/s12889-020-10007-w

77 Stassen W, Wallis L, Vincent-Lambert C, et al. The proportion of South Africans living within 60 and 120 minutes of a percutaneous coronary intervention facility. Cardiovascular Journal of Africa 2018;29:6–11. doi:10.5830/CVJA-2018-004

78 Nyandwi E, Veldkamp A, Amer S, et al. Schistosomiasis mansoni incidence data in Rwanda can improve prevalence assessments, by providing high-resolution hotspot and risk factors identification. BMC Public Health 2017;17. doi:10.1186/s12889-017-4816-4

79 Manongi R, Mtei F, Mtove G, et al. Inpatient child mortality by travel time to hospital in a rural area of Tanzania. Tropical Medicine and International Health 2014;19:555–62. doi:10.1111/TMI.12294

80 Ansumana R, Malanoski AP, Bockarie AS, et al. Enabling methods for community health mapping in developing countries. International Journal of Health Geographics 2010;9. doi:10.1186/1476-072X-9-56

81 Oteri J, Idi Hussaini M, Bawa S, et al. Application of the Geographic Information System (GIS) in immunisation service delivery; its use in the 2017/2018 measles vaccination campaign in Nigeria. Vaccine 2021;39:C29–37. doi:10.1016/j.vaccine.2021.01.021

82 Borgdorff MW, A Walker GJ. Estimating vaccination coverage: Routine information or sample survey? J Trop Med Hyg 1988;91:35–42.

83 Barker RD, Nthangeni ME, Millard FJC. Is the distance a patient lives from hospital a risk factor for death from tuberculosis in rural South Africa? International Journal of Tuberculosis and Lung Disease 2002;6:98–103.

84 Clur S-A. Frequency and severity of rheumatic heart disease in the catchment area of Gauteng hospitals, 1993-1995. South African Medical Journal 2006;96:233–7.

85 Iyun F. Hospital service areas in Ibadan city. Soc Sci Med 1983;17:601–16.

86 Okiro EA, Alegana VA, Noor AM, et al. Malaria paediatric hospitalization between 1999 and 2008 across Kenya. BMC Med 2009;7:75. doi:10.1186/1741-7015-7-75

87 Okiring J, Routledge I, Epstein A, et al. Associations between environmental covariates and temporal changes in malaria incidence in high transmission settings of Uganda: a distributed lag nonlinear analysis. BMC Public Health 2021;21. doi:10.1186/s12889-021-11949-5

88 Stewart K, Li M, Xia Z, et al. Modeling spatial access to cervical cancer screening services in Ondo State, Nigeria. International Journal of Health Geographics 2020;19. doi:10.1186/s12942-020-00222-4

89 Wilson DP, Blower S. How far will we need to go to reach HIV-infected people in rural South Africa? BMC Medicine 2007;5. doi:10.1186/1741-7015-5-16

90 Nelli L, Guelbeogo M, Ferguson HM, et al. Distance sampling for epidemiology: An interactive tool for estimating under-reporting of cases from clinic data. International Journal of Health Geographics 2020;19. doi:10.1186/s12942-020-00209-1

91 Ouma P, Macharia PM, Okiro E, et al. Methods of Measuring Spatial Accessibility to Health Care in Uganda. 2021. 77–90. doi:10.1007/978-3-030-63471-1_6

92 Delmelle EM, Desjardins MR, Jung P, et al. Uncertainty in geospatial health: challenges and opportunities ahead. Annals of Epidemiology. 2022;65. doi:10.1016/j.annepidem.2021.10.002

93 Herbreteau V, Salem G, Souris M, et al. Thirty years of use and improvement of remote sensing, applied to epidemiology: From early promises to lasting frustration. Health and Place 2007;13. doi:10.1016/j.healthplace.2006.03.003

94 Nachmany Y, Alemohammad H. Detecting roads from satellite imagery in the developing world. In: IEEE Computer Society Conference on Computer Vision and Pattern Recognition Workshops. 2019.

95 Banke-Thomas A, Macharia PM, Makanga PT, et al. Leveraging big data for improving the estimation of close to reality travel time to obstetric emergency services in urban low- and middle-income settings. Frontiers in Public Health 2022;10. doi:10.3389/fpubh.2022.931401

96 Woods D, Cunningham A, Utazi CE, et al. Exploring methods for mapping seasonal population changes using mobile phone data. Humanities and Social Sciences Communications 2022;9:247. doi:10.1057/s41599-022-01256-8

97 Gesicho MB, Were MC, Babic A. Data cleaning process for HIV-indicator data extracted from DHIS2 national reporting system: a case study of Kenya. BMC Medical Informatics and Decision Making 2020;20. doi:10.1186/s12911-020-01315-7

98 Herringer M. Health catchment areas. 2021.https://github.com/healthsites/healthsites/wiki/Health-catchment-areas (accessed 3 Aug 2022).

